# Altered resting-state network dynamics in Schizophrenia

**DOI:** 10.1101/2020.07.21.20157347

**Authors:** Ana R. Lopes, Anna Letournel, Joana Cabral

## Abstract

Schizophrenia remains a poorly understood disease, hence the interest in assessing and indirectly characterizing brain activity and connectivity. This paper aims to search for potential biomarkers in schizophrenia with functional magnetic resonance data, between subjects in the resting state. Firstly, we used fMRI from an open database, SchizConnect, of 48 subjects, in which 27 were control subjects, with no apparent disease and the others 21 were patients with schizophrenia. With the SPM tool, we proceeded to manually pre-process the images obtained, at the risk of having influenced the final results. Then, with the AAL atlas as a reference, we divided the brain into 116 areas. Then, brain activity in these areas were analysed, using the LEiDA method, which aims to characterize brain activity at each time point t by phase locking patterns of the BOLD signal. After the application of LEiDA, brain activity was evaluated based on trajectories and bar graphs of functional connectivity states in which the probability of occurrence and their dwell time were calculated for each state. It was also found that the visual cortex was the subsystem that showed significantly more probability of occurrence in schizophrenia patients to be assessed, and may correspond to symptoms of hallucinations by the patients with schizophrenia.

## 1. Introduction

Schizophrenia currently affects 20 million people ^[1]^, with prevalence in young male adults ^[2]^. Although it is not as prevalent as other psychiatric illnesses, it is associated with various physical diseases, such as cardiovascular, metabolic and infectious diseases, which can affect mobility and behaviour ^[1]^.

The preferred technique in this work is to measure activity of fMRI, using the BOLD signal. Functional connectivity is characterized by spatial patterns that form over time and is generally measured as Pearson’s correlation between the BOLD signals over time. The phase coherence connectivity of the BOLD signal is used to obtain a dynamic FC matrix ^[3]^.

This paper involved the research for a potential biomarker in schizophrenia. If the possibility those successful results are to be obtained, it will be necessary to pursue more specific studies and demonstrate consistent results that can be validated and used as a clinical biomarker in the future ^[4]^.

The analysed images were taken from an open database, SchizConnect.

We selected a manual division of the three-dimensional image of the brain into slices, in order to analyse them individually.

The LEiDA method is very robust to high frequency noise, which affect the almost instantaneous measurements of FC when exceeding a certain limit. The defined spatial and temporal scales are the determining factors for LEiDA to be able to detect the number and shape of each FC pattern ^[3]^.

## 2. Methods

### 2.1. Image Acquisition

We used fMRI data from patients with schizophrenia and matched controls from the SchizConnect (http://schizconnect.org/) database of neuroimaging of schizophrenia patients ^[5]^.

The images were obtained using the 3T (Tesla) Siemens, with a repetition time (TR) of 2 seconds and with an echo time (TE) of 29 milliseconds.

### 2.2. subjects

Due to the high amount of data existing for fMRI, we chose to restrict the data to subjects between 18 and 30 years of age and, when obtaining the functional magnetic resonance images in resting state.

By reducing the number of data, we obtained 48 subjects, of which 27 were part of the control group, with no associated pathology and the remaining 21 were patients with schizophrenia. When carrying out the statistical evaluation using ttest2, we did not find significant differences between the ages of the subjects, since we obtained a value of p=0.31204 and to verify differences, we would have to obtain a value smaller then 0.05.

**Table 1.** Characteristics of the subjects.

|  | Control group | Group with schizophrenia |
| --- | --- | --- |
| Age | $25 \pm 2,7$ | $24 \pm 3,4$ |
| Genre | 18 men, 9 women | 19 men, 2 women |

### 2.3 Pre-processing

The data was pre-processed using a statistical parametric mapping software, SPM12 ^[6]^ designed to work with MATLAB ^[7]^, in which the format is NIFTI-1 (.nii).

These were obtained with an original acquisition size of 79 × 95 × 79 × T (X × Y × Z × T) and subsequently changed to correspond to the MNI152 space ^[8]^, into a size of 91 × 109 × 91 × T.

Initially, the Realign: Estimate function was used, which aims to eliminate movement artifacts and align the series of acquired volumes for functional analysis. After being realigned, the Normalize: Estimate & Write function was applied, normalizing the data for the MNI space, which is an essential step for the use of atlases in the process in which the subject’s volumes are transformed in order to adjusted to the template of the MNI space. After normalization, only one image of each subject was selected, in image to align, to serve as a spatial reference. This served to spatially alter all the other images of each subject, in images to write, all the subject’s images were placed in that reference space, resulting in the end, in an alignment of all images.

In order to increase the signal-to-noise ratio, the Smooth function was used, using an 8mm Gaussian filter for the images obtained.

### 2.4. Parcellation

It can be considered that parcellation subdivide the cortex into a set of specific homogeneous regions of the subject himself. To make the parcellation we used the code ParcelsMNI2mm ^[9]^.

In order to reduce the dimensionality of the data (voxel × t), the AAL atlas was used to define the brain into 116 cortical and subcortical regions, including the cerebellum. The data were reduced to a size N × t, where t = 150 TR, performing an average of the BOLD signal of all the voxels associated with each region of the brain.

### 2.5. Statistical Analyses

LEiDA was applied to capture FC patterns from fMRI data in a single TR resolution with reduced dimensionality. In a first phase, the BOLD signals in the brain areas, using the AAL atlas ^[10]^ were filtered and subsequently the phase of the filtered BOLD signals was estimated using the Hilbert transform (**Equation 1**).

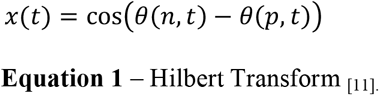

Given the BOLD phases, a dynamic FC matrix was calculated based on the BOLD phase coherence, where each dynamic FC captures the degree of synchronization between areas n and p at time t (**Equation 2**) ^[12]^. The phase locking matrix of dFC (n, p, t), which estimates the orientation of the phase locking between brain areas of each pair of nodes at time t, can be determined by calculating the cosine of the phase difference, with the following formula ^[13]^:

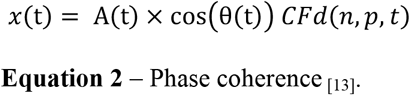

The leading eigenvector of the N × N phase locking matrix at time t is an N × 1 vector, which captures the main orientation of the BOLD phases in all areas, where each element represents the projection of the BOLD phase in each brain area for the main eigenvector ^[14]^.

In order to be able to describe the average time in a given state of phase locking, in each fMRI scan, the dwell time is calculated. The same (**Equation 3**) is defined by 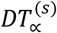 and expresses the average of all periods in each state, which are characterized as ∝. It also lists the value of *P*_∝_, defined as the number of consecutive periods for the state to be evaluated and 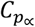 as the duration of each of these consecutive periods ^[11]^.

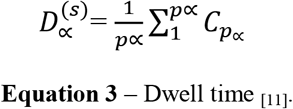

The k-means algorithm was used, which is characterized as an interactive process, which aims to minimize the distance of each observation and the nearest centroid. This algorithm is used to group the main eigenvectors in clusters from k = 2 to k = 20, obtaining a total of 19 partitions ^[12]^. Each calculation was performed 300 times to ensure consistency in the results.

## 3. Results

### 3.1. Detection of recurring BOLD phase locking patterns

LEiDA is only capable of detecting phase locking patterns, so variations in amplitude will not have any influence, and changes in signal are not sensitive to variations in amplitude ^[12].^

In figure 2, a relatively high start of acquisition of the BOLD signal is observed, which may correspond to artifacts. However, with the method used, the signal information is collected for each t only after the second TR, so the initial values will not have any influence on the data obtained.

**Figure 1.**
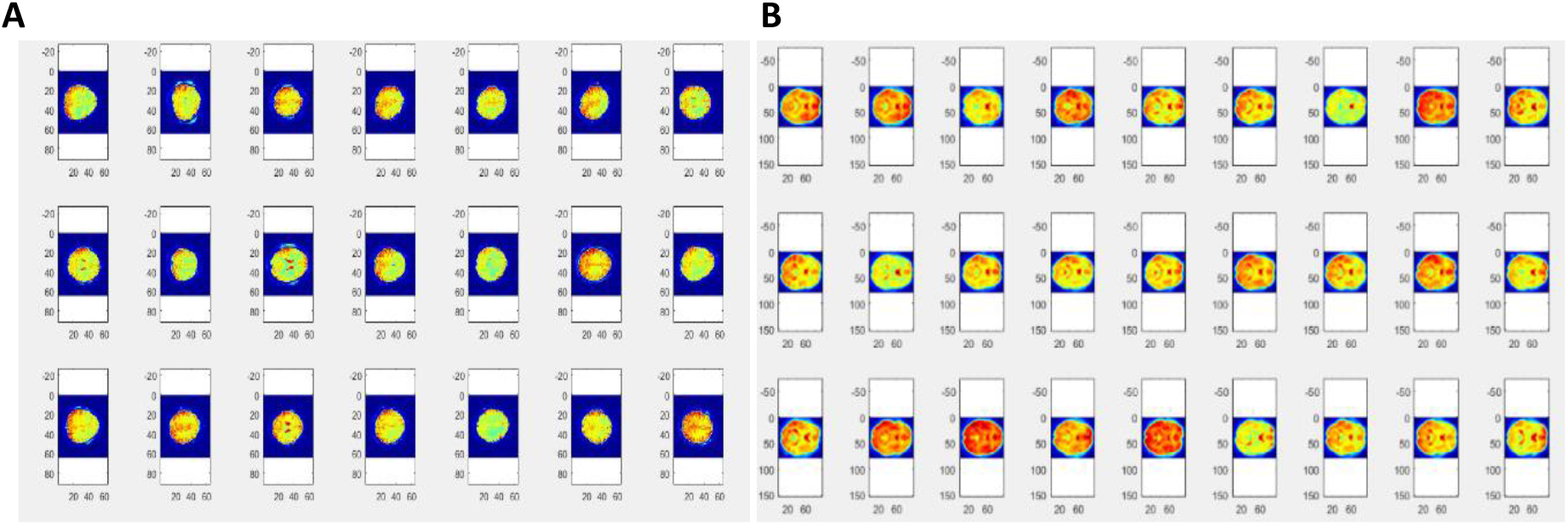
Images of brains from subjects with schizophrenia. (**A**) Image of the brains in the original space. (**B**) Image of the subjects’ brains after pre-processing to the MNI space.

**Figure 2.**
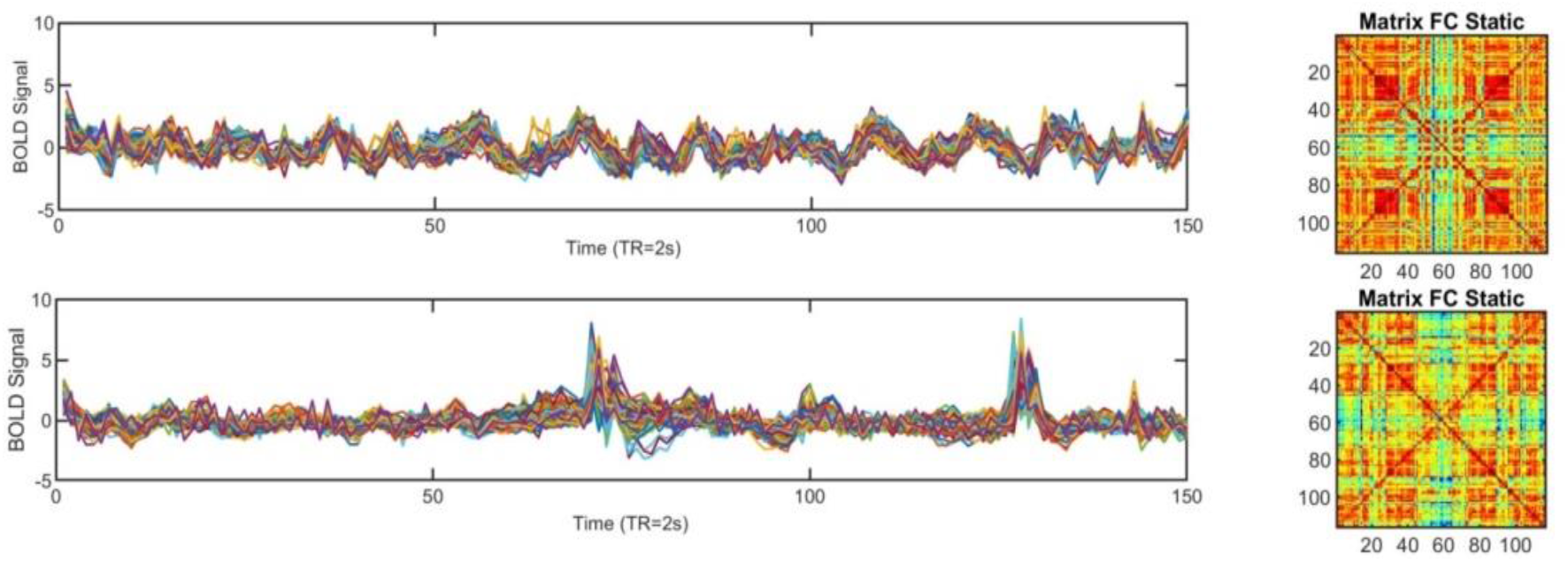
Variation of BOLD signal in one subject. (**A**) Representative subject of the control group and (on the right) their respective static FC matrix. (**B**) Representative subject of the group with schizophrenia and (on the right) their respective static FC matrix.

Since LEiDA focuses on BOLD phase locking patterns, the existence of movement artifacts only affects the occurrence of global synchronization patterns. Thus, in order to minimize the consequences of removing these artifacts on the final results, this method was analysed directly on the uncorrected BOLD signal. The results can then be analysed taking into account the degree of movement of each subject in the scan (which was not provided in the present database). Comparisons between the variations obtained can be seen in figure 2A.

The static FC matrix does not allow the visualization of the evolution over the states ^[3]^. In this way, a number of clusters is selected, k=10 (as k can vary from 2 to 20, k = 10 was chosen, for being a number in the middle), which allows the observation of evolution with 10 different matrices (figure 3).

**Figure 3.**
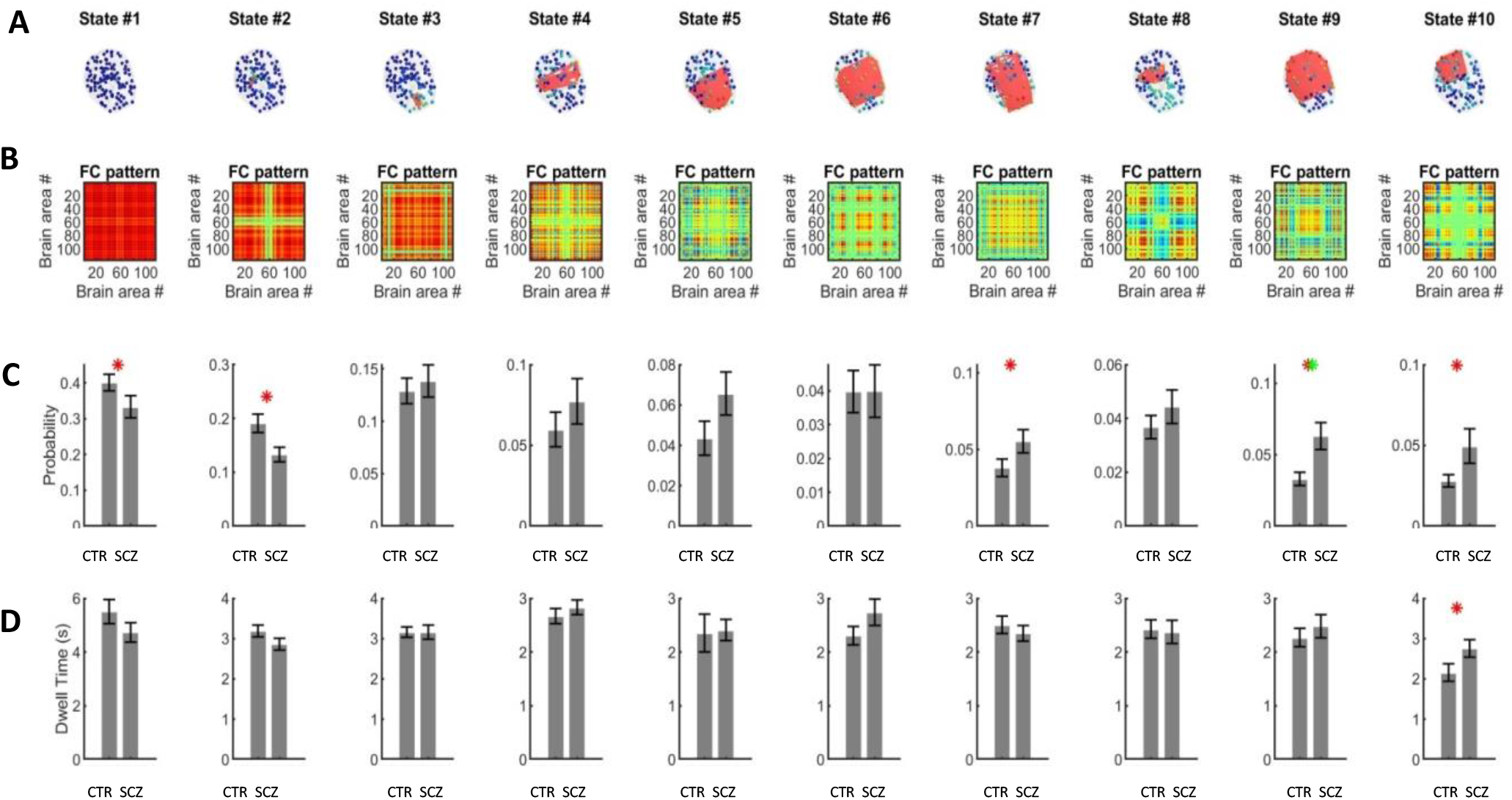
Visualization of different forms of the affected areas and the respective matrices for each state for k = 10, in resting state and associated standard deviation. (**A**) A network in cortical space, where the value of VC (n) is used to scale the color of each brain area and links are plotted between areas to highlight the network detaching from the global mode. (**B**) A matrix obtained by calculating the outer product of VC, where positive values are the product of VC elements with the same sign, be they positive or negative. (**C**) Probability of occurrence (mean standard error of the mean across subjects) of each PL state. (**D**) Probability of dwell time (mean standard error of the mean across subjects) of each PL state. CTR = Control; SCZ = Subjects with schizophrenia.

Therefore, this figure allows the visualization of the results obtained in different ways, in which in the upper zone (figure 3A), a transparent brain is analysed for each standard state, the same corresponding to the MNI152 template ^[8]^.

The red lines reflect the connections between the areas whose BOLD signal is displaced ^[15].^ Analysing the matrices, was verified the existence of different patterns, being thus quite visual, not only at the level of the brain but also in the respective matrices for each state.

These images allow us to see a bigger picture since both the observation of the affected areas in the brain by the networks and they also provide the visualization of the matrices coinciding with each state (figure 3B).

After the characterization of the patterns, there is a transition to a resting state that reduces all samples to the number of selected clusters, which will correspond to the sum of these same patterns. The weighted sum of these matrices corresponds to the static FC matrix.

The bar graphs shown in the lower part (figure 3D) are related to each state. These help in the comparative assessment of probability of occurrence and dwell time in each state, between subjects with schizophrenia and control subjects.

We can visualize an increased probability of finding the control subjects in relation to the subjects with schizophrenia in the global state (PL1). In this study, another global state (PL2) is assessed, where there is also a greater probability of finding control subjects compared to subjects with schizophrenia.

In the bar graph, it is possible to observe red and green asterisks, while red ones can correspond to false positives, the green asterisk appears since the data shows greater consistency, which can correspond to a true positive. The global state can be defined as stability, the capacity to inhibit and ensure that the locking phase does not shift excessively to other networks.

In figure 4, PL1 can be considered to be a global state and, since PL2 state is similar to PL1, it can also be characterized as a global state.

**Figure 4.**
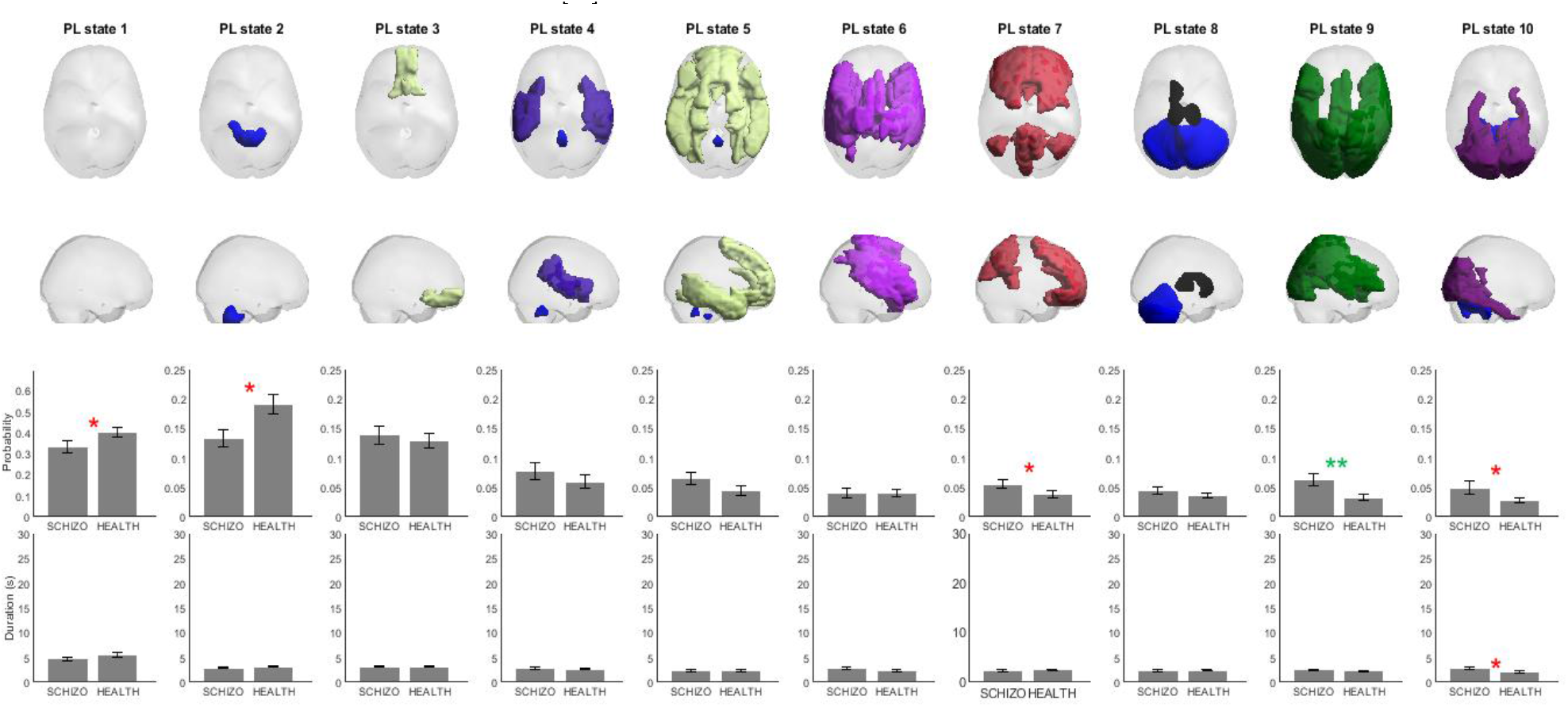
Interpretation of all brain areas with positive values in the vector, colored according to the functional network to which they show a greater overlap at resting state for k = 10.

This type of comparative assessment makes it possible to differentiate networks that show changes from those that do not.

To divide the brain into the 7 functional networks, according to the reference given by Yeo ^[16]^, each brain network will correspond to a specific color. Were the violet color corresponds to the visual network, the dark blue color to the somatosensory network, the green color to the dorsal attention, the pink color to the ventral attention, the cream color to the limbic, the orange to the frontoparietal and the red to the default mode network ^[16]^.

In figure 4B, subjects with schizophrenia have much higher values compared to control subjects in all states, with the exception of the global ones. The most significant states are PL1, PL2, PL7, PL9 and PL10, since their probability of occurrence changes significantly between groups, surviving correction for multiple comparisons for state 9 (green asterisks). It is also analysed a consistency in the affected networks, visual, default mode network and dorsal attention.

The probability of the PL6 state occurring is approximately equal for both groups.

In order to color the brain regions in figure 4, the Yeo reference mask is used ^[16]^.

When evaluating the dwell time relative to each state, there are no major differences, just the appearance of an asterisk in order to report significance in PL10. This figure also shows a black zone, a baseline activation network, in the PL8, which is higher in subjects with schizophrenia.

### 3.2. Detection of the different functional connectivity states

To evaluate the p values associated with the comparison between all the probabilities of the states and their dwell times, three limits are characterized, being observed with the colors red, green and blue. These limits are identified as points above the 0.05 threshold, with black color (not significant) and below this threshold, clusters are red (may represent false positives) ^[14].^

As shown in the figure, asterisks with the colors corresponding to the Yeo reference (figure 5) were placed in the centroid that exceed the probability limits *p* <0.05 / *k* and p<0.05 / Σ(*k*). Both limits are shown in dashed lines, in green and blue, respectively.

**Figure 5.**
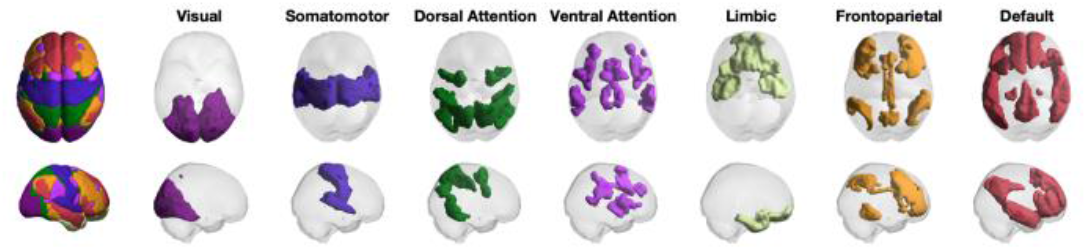
Yeo reference networks ^[16].^

For an evaluation of all centroids in all states, from k=2 to k=20, these figures were created. However, the centroids corresponding to k=1 are not present in them, since it is always considered as a global state and therefore the probability of occurrence between states and the dwell time in each of them is not evaluated.

By analysing the figure, it is understood that the affected area was the visual area, since the inserted brains are all the same color (figure 6), which corresponds to the visual area by Yeo’s reference (figure 5).

**Figure 6.**
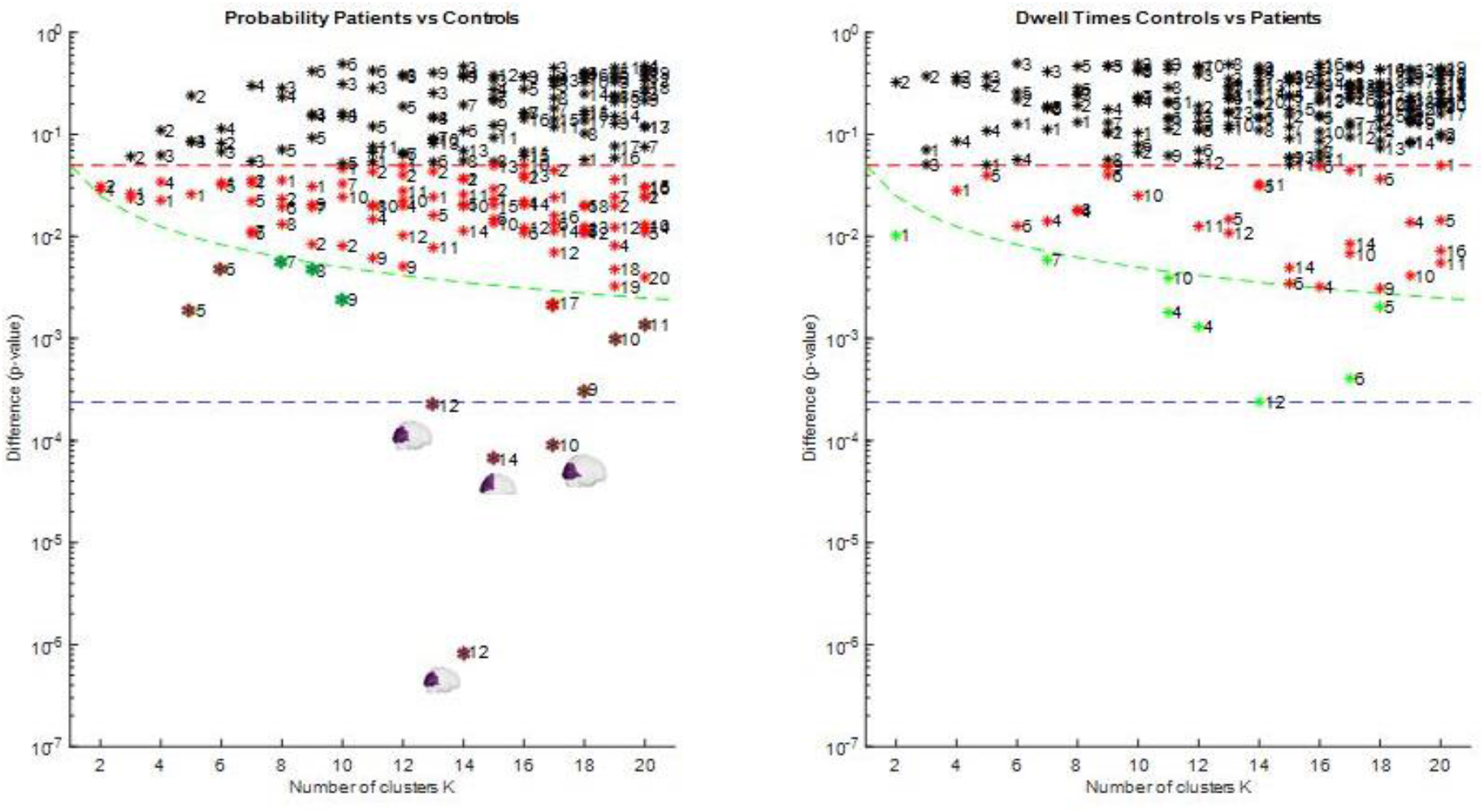
Interpretation of the p values, for the probability of occurrence between states (left) and for the probability of dwell time at each state (right).

This figure shows a very significant increase in terms of the probability of occurrence between states, since there are four values below the blue limit. Of these four, the value corresponding to k=13 and C=12 and the value of k=14 and C=12 are even lower.

The clusters and their affected brain areas are also evaluated, predominantly in the visual areas, but also in the dorsal and ventral attention networks, in figure 7. More representative values can be seen, which correspond to k=13 and C=12, k=14 and C=12, k=15 and C=14 and also to k=17 and C=10.

**Figure 7.**
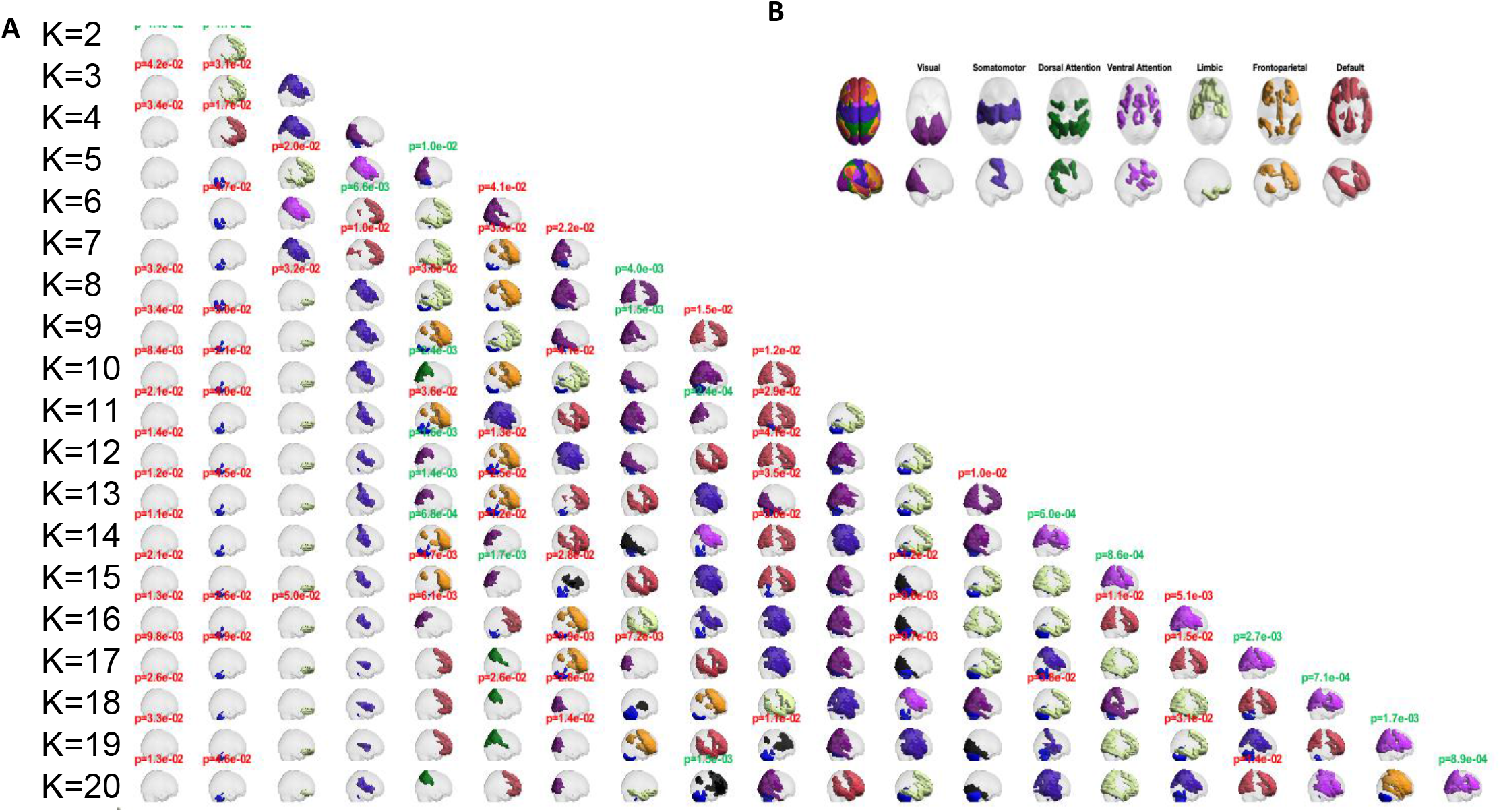
Pyramid with the centroid obtained from LEiDA, colored according to the Yeo reference networks ^[16].^ (A) Pyramid with centroids obtained from LEiDA from k = 2 to k = 20. (B) Yeo reference networks ^[16]^.

In figure 8, these states are observed separately in order to be able to compare them statistically.

**Figure 8.**
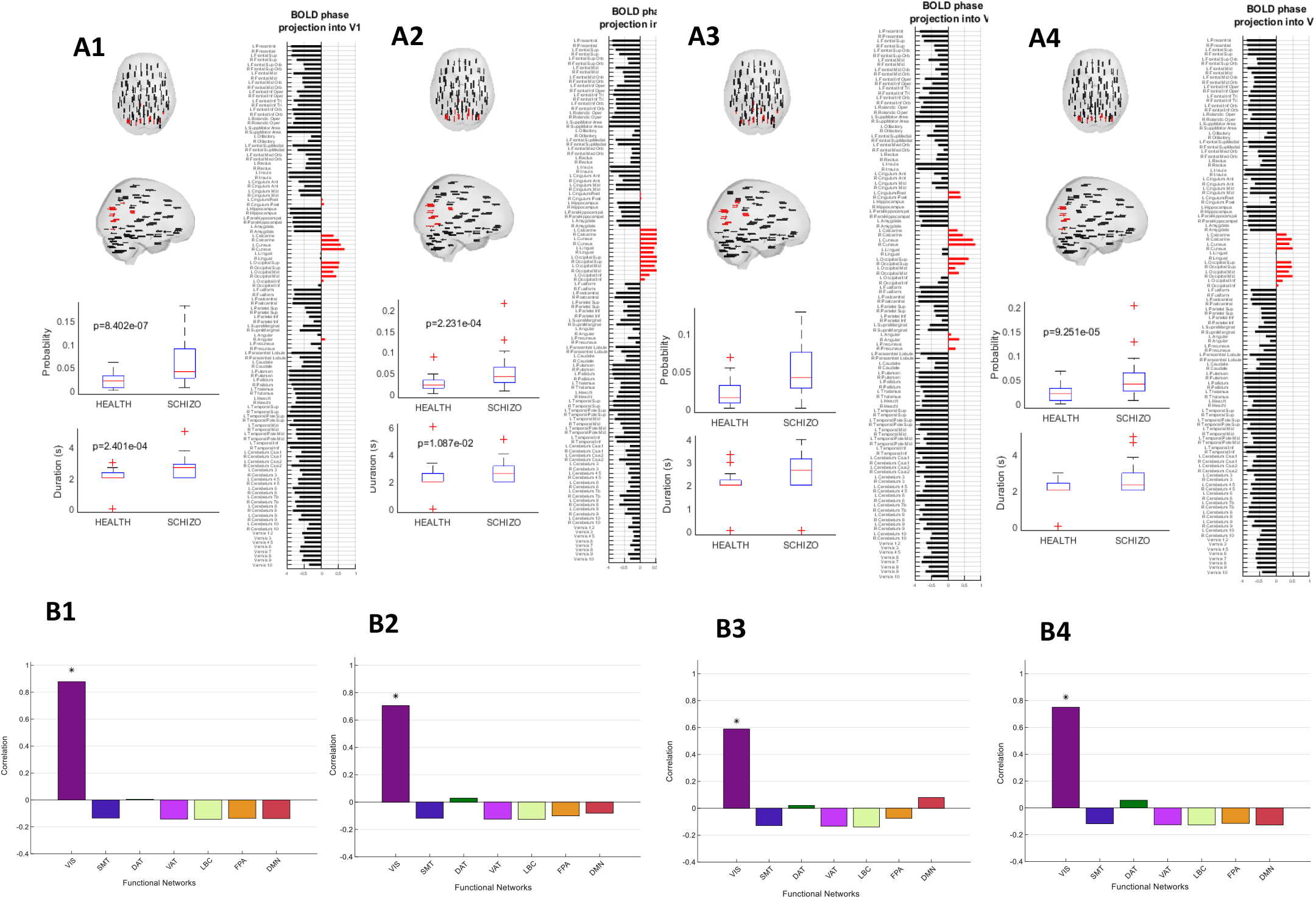
Patterns of BOLD phase locking exhibiting significant differences in probability of occurrence between groups. (**A1**) Evaluation of the direction of the vector in the standard brain, the bar graph corresponding to the phase projection of the BOLD signal for k=14, C=12 and the graph with the probability of occurrence and the dwell time there. (**A2**) Evaluation of the direction of the vector in the standard brain, the bar graph corresponding to the phase projection of the BOLD signal for k=13, C=12 and the graph with the probability of occurrence and dwell time in it. (**A3**) Evaluation of the direction of the vector in the standard brain, the bar graph corresponding to the phase projection of the BOLD signal for k=15, C=14 and the graph with the probability of occurrence and the dwell time in it. (**A4**) Evaluation of the direction of the vector in the standard brain, the bar graph corresponding to the phase projection of the BOLD signal for k=17, C=10 and the graph with the probability of occurrence and dwell time in it. (**B1**) Correlation of the A1 centroid based on the Yeo reference networks ^[16].^ (**B2**) Correlation of the A2 centroid based on Yeo reference networks ^[16].^ (**B3**) Correlation of the A3 centroid based on the Yeo reference networks ^[16].^ (**B4**) Correlation of the A4 centroid based on the Yeo reference networks ^[16]^.

### 3.3. Most relevant FC states

The most relevant values are evaluated separately, with the bar graphs verifying the partitions of the 116 areas, in figures 8 A1 to A4 that correspond to the projections of the BOLD signal locking phase in each of the vectors.

When comparing the figure from A1 to A4, it is possible to observe the areas in which the phase projection of the BOLD signal is moving to the visual areas of the AAL atlas (bars in red). The same is also true when comparing the brains with the arrows, located in red, in that area.

It should be noted that in figure 8 B1, which corresponds to the most significant centroid, there is no correlation with any other network of the value in question. Something that is not observed in the other 3 centroids to be evaluated, since they show a low correlation between dorsal attention and the default mode network.

In figure 8 B2 and figure 8 B4, there is a correlation of the visual network with the dorsal attention network, and in figure 8 B3, a correlation of the visual network with the dorsal attention network, but with one more coactive network, the default mode network.

## 4. Discussion

Schizophrenia does not affect only one area, it affects contributions among several, in which each can lead to different symptoms, taking into account that people with schizophrenia will have different representations of the disease. Such observation may correspond to the results obtained, in which there was a correlation between brain areas, which may have different contributions to the symptoms that the patient expresses, and we validate this theory by obtaining a correlation between the visual network with the dorsal attention network, the frontoparietal and the default mode network.

For comparison with studies carried out with parcellation, in which the areas corresponding to the cerebellum were excluded, it is observed that a “new” network appears, of black color considered basal, which may inhibit the activation of the other networks that are not needed. The presence of this can be seen due to the 116 brain areas being considered. When comparing with studies carried out, there was a change in the PL2 state, since, in the present study, it also corresponds to a global state.

The static matrix is defined as the weighted sum of all other matrices and, although it seems that this sum is performed simultaneously, what really happens is a constant evolution of the matrix. When executing the k-means clustering method, the cluster intends to characterize the patterns, moving to a space state in which the samples are reduced to a defined number of clusters, which represent the total sum.

A curiosity about ghost attractors is the difficulty of being able to detect them visually, since they occur in a TR. However, if they are recurrent and these results are constantly appearing, they may correspond to something relevant for study. Since the TR used was 2 seconds is a probabilistic question as to whether or not this signal can be captured, since it must be at the same time that the MRI spin is sent. These states demonstrate a short period of life, being faster than the BOLD signal, hence the inherent difficulty in observing and characterizing them temporally. Therefore, with the probabilistic assessment, it is possible to calculate the probabilities associated with its occurrence. This time corresponds to an interpretation of the dwell time, being the same, always higher for the global state in which it has more stability while the others are unstable.

Reference networks were defined using a specific method, which is used as a model without actually having a standard. These networks are not considered perfect, since the method used to obtain them is based on correlations. The echoes present in the resting state allow the observation of the brain structure, with the possibility that these and the dorsal attention network, which increases in this disease, are responsible for the hallucinations caused in patients with schizophrenia.

The realization of echo may be related, perhaps, to the positive probability value of the default mode network. Among these subjects there is variability, and it is possible that some of them only have visual or sensory hallucinations and when there is an involvement of the dorsal attention network, it could originate another type of symptoms.

The statistical approach we used was exactly the same for all subjects. It does not correct artifacts, since their correction is never ideal and, when trying to correct them, more artifacts may originate in pre-processing or perhaps even correct essential signals. As we did not make this correction, the results obtained may or may not correspond to the reality of the data.

### 4.1. Limitations

One of the limitations of this paper is related to the mask used not being in accordance with the space to register the signals. In this way we carried out an adaptation of the patients’ brains manually, thus obtaining less precise and cohesive results, where the overlapping networks are not symmetrical. These could be improved if we had chosen to use our own tools for that effect in parcellation.

However, even with the use of other tools, the overlap with the Yeo reference networks or with the template MNI 152 would never be perfect, since the brains are quite different from each other.

## Data Availability

All data is from SchizConnect

http://schizconnect.org/

## Acknowledgments

Data used in preparation of this article were obtained from the SchizConnect database (http://schizconnect.org) As such, the investigators within SchizConnect contributed to the design and implementation of SchizConnect and/or provided data but did not participate in analysis or writing of this report.

Data collection and sharing for this project was funded by NIMH cooperative agreement 1U01 MH097435.

## Notes

### Competing Interest Statement

The authors have declared no competing interest.

